# Critical appraisal of the European Union Scientific Committee on Health, Environmental and Emerging Risks (SCHEER) *Preliminary Opinion on electronic cigarettes*

**DOI:** 10.1101/2020.11.09.20228296

**Authors:** R. O’Leary, R. Polosa, G. Li Volti, on behalf of the CoEHAR study group

## Abstract

**Background:** In preparation for the 2021 revision of the European Union Tobacco Products Directive, the Scientific Committee on Health, Environmental and Emerging Risks (SCHEER) has posted its *Preliminary Opinion on Electronic Cigarettes*. They conclude that e-cigarettes only achieve a sub-optimal level of protection of human health. In this paper we provide evidence that *Opinion’s* conclusions are not adequately backed up by scientific evidence and totally disregard the potential health benefits of using alternative combustion-free nicotine containing products as substitute for tobacco cigarettes.

**Method:** Searches for articles were conducted in PubMed and by citation chasing in Google Scholar. Articles were also retrieved with a review of references in major publications. Primary data from World Health Organization surveys, the conclusions of reviews, and peer-reviewed non-industry studies were cited to address errors and omissions identified in the *Opinion*.

**Results:** The *Opinion* omitted reporting on the individual and population health benefits of the substitution of e-cigarettes (ENDS) for cigarette smoking. Alternative hypotheses to the gateway theory were not evaluated. Its assessment of cardiovascular risk is contradicted by numerous reviews. It exhibits biases in its statements from the measurements selected on the frequency of use. It did not report non-nicotine use. It misrepresented trends in ENDS prevalence. It over-emphasized the role of flavours in youth ENDS initiation. It did not discuss cessation in sufficient length.

**Conclusions:** For the delivery of a robust and comprehensive final report, the members of the Working Group of the Scientific Committee on Health, Environmental and Emerging Risks will need to consider (1) the potential health benefits of ENDS substitution for cigarette smoking, (2) alternative hypotheses and contradictory studies on the gateway effect, (3) its assessment of cardiovascular risk, (4) biases arising from the measurements of frequency of use, (5) non-nicotine use, (6) the role of flavours, and (7) a fulsome discussion of cessation.

## Background

The European Union (EU) is in the process of revising the Tobacco Products Directive Article 28 to be submitted to the EU Parliament by 20 May 2021. The Scientific Committee on Health, Environmental and Emerging Risks (SCHEER) “Request for a scientific Opinion on Electronic cigarettes” [1] was mandated on February 7, 2019, and the *Preliminary Opinion on electronic cigarettes* [2] was posted online on September 23, 2020. An online comment process allowed a one-month period for the submission of comments, with a limit of 3800 characters for each section of the *Opinion*. The Center of Excellence for the Acceleration of Harm Reduction (CoEHAR) at the University of Catania, Italy approved the submission of comments on the *Opinion*. This paper greatly expands on the comments submitted to the online comments process and cites many additional studies and reviews. We refer to e-cigarettes as ENDS (electronic nicotine delivery systems), the terminology used by the World Health Organization (WHO). Their terminology could also be applied to another popular and very different product category, heat-not-burn or “heated tobacco product” that is electronic and delivers nicotine, but we have not included those products in this paper.

In reading the *Preliminary Opinion*, we were struck by its omission of any discussion of tobacco harm reduction, its blindered focus on the disputed gateway effect of youth ENDS use on cigarette initiation, and its unjustified appraisal of cardiovascular risks. A closer reading revealed its biases in reporting the frequency of ENDS use, its misrepresentation of the role of flavours, and its omission of reporting on non-nicotine use.

Our critical appraisal of the *Opinion* was sharpened by the comments of other individuals and organizations. Dr. Karl E. Lund at the Norwegian Institute of Public Health circulated comments and references within an email network. Clive Bates posted two critiques [3] [4]. Christopher Snowdon at the European Policy Information Center (EpiCenter) posted a response [5]. These publications provide additional criticisms of the *Opinion*.

Our critique seeks to address the errors and omissions we found in the *Preliminary Opinion*. Our critical appraisal is based on evidence from primary data (the WHO in particular), the conclusions of systematic and narrative reviews, and the findings of studies conducted by non-industry researchers.

## Methods

Multiple searches were conducted to obtain the primary data, reviews, and studies to support the critical appraisal of the *Opinion*. In addition to the searches, we referenced two authoritative reviews on ENDS.

We used three primary sources of data. To address the *Opinion’s* statements on renormalization and a possible impact of ENDS use on quit trends we extracted primary data from the most recent WHO reports [6] [7]. For the prevalence of ENDS use we extracted data from the Passport database by EUROMONITOR [8], an established consumer products research organization.

We searched PubMed for reviews and studies on the subject areas of cessation, cardiovascular risks, the gateway effect, and the impact of flavours on youth initiation and adult cessation. Searches were conducted with the terms “e-cigarettes” AND “review” AND [subject keyword] in the title/abstract field. Subject keywords: cessation, cardiovascular, youth (OR student OR adolescent), biomarkers, prevalence. These searches were performed individually for each subject keyword. Another set of searches was conducted with the keywords “e-cigarette” AND [individual EU member state] in the title/abstract field.

Studies and reviews were also identified from published authoritative key sources including the *Public Health Consequences of E-Cigarettes* [9] and the *Report on the Scientific Basis of Tobacco Product Regulation: Seventh Report of a WHO Study Group* [10]. We also referenced studies from *Vapour Products/E-Cigarettes: Claims and Evidence* [11] and the monograph *Clearing the Air: A systematic review on the harms and benefits of e-cigarettes and vapour devices* [12]. Finally, reviews and studies in the *Opinion* were searched for additional findings. All reviews and studies retrieved were citation searched (snowball search) in Google Scholar to identify additional publications.

The results of the search were reviewed for best fit for our comments on the *Opinion* as the comment sections were strictly limited in size. Studies of EU member states were prioritized over those from the United States (US) due to the substantial differences between their ENDS regulations and their commercial markets. Nevertheless, for some subjects only US research was available or provided major studies. These data, reviews, and studies were used to address the omissions and errors we observed in the *Opinion*, discussed in the critical appraisal that follows.

### Omitted from the Terms of Reference: Tobacco Harm Reduction

The background section of the *Request for Scientific Opinion* included the role of ENDS in harm reduction. Yet the evaluation of harm reduction was not specifically stated as an item in the Terms of Reference, obliquely referring to “their use.” This vague statement in the *Request* resulted, intentionally or unintentionally, in the omission of the key role of ENDS in tobacco harm reduction. The potential effects of ENDS on individual and public health is the subject of a tremendous number of studies and medical association position statements. The *Opinion* is seriously incomplete without an evaluation of ENDS for tobacco harm reduction. We present evidence on how ENDS have the potential to reduce the risks of smoking for the substantial number of EU citizens who smoke.

#### Not Quitting

The assumption that people who smoke want to quit is drawn from self-reports in surveys, but it constitutes a vague wish. On the other hand, when asked directly about quitting activities, a staggeringly high number of EU adults who smoke have no intention to quit [13].

Furthermore, for those that wish to quit, cigarette quit rates are very low, from 3%–12%, and relapse rates are very high, from 75%–80% in the first six months and 30%– 40% even after one year of abstinence [see studies cited in 14]. Quitting is not a single event but a dynamic process, and relapse is a common component of this process. While international guidelines place great emphasis on relapse prevention, very little can be offered to help smokers who have relapsed [15]. This challenge calls for innovative and effective alternatives for relapse prevention, including the use of ENDS.

#### Reduced Emissions Compared to Cigarettes

For those who do not make the effort to quit, and for those who cannot quit or remain abstinent, ENDS may provide an alternative to cigarettes. It is generally accepted that ENDS have substantially lower and fewer toxic emissions than cigarettes. The National Academies of Science, Engineering, and Medicine state “There is substantial evidence that except for nicotine, under typical conditions of use, exposure to potentially toxic substances from e-cigarettes is significantly lower compared with combustible tobacco cigarettes” [9, p. 6]. For example, emissions testing [16] found that ENDS toxicant emissions are 9 to 450 times lower than cigarette smoke. Furthermore, many toxic substances in cigarettes are not emitted by ENDS, as clearly illustrated in the research [17] that did not detect 61 of 79 compounds present in tobacco smoke (tally of Table 1). These findings are corroborated in the review by the European Respiratory Society [18] and other reviews [19, 20]. Stephens [21] calculated the cancer potency of ENDS emissions to have 0.004 of the relative lifetime cancer risk of tobacco smoke.

**Table 1.**
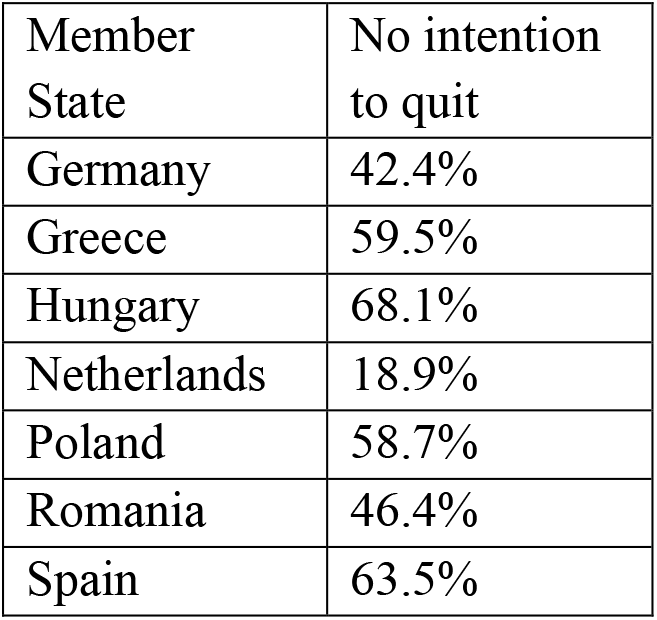
Prevalence of EU Adults with No Quit Intentions Source: [13]

| Member State | No intention to quit |
| --- | --- |
| Germany | 42.4% |
| Greece | 59.5% |
| Hungary | 68.1% |
| Netherlands | 18.9% |
| Poland | 58.7% |
| Romania | 46.4% |
| Spain | 63.5% |

Source: [13]

While major assumptions and guesswork are required to translate reductions in emissions into an estimate of actual health risk, it is impossible to believe that the orders-of-magnitude differences do not represent enormously lower risk. The *Opinion* never once compares the emission levels of ENDS vapour with cigarette smoke.

The UK regulatory organization, Committee on Toxicity of Chemicals in Food, Consumer Products and the Environment conducted an extensive and systematic review on the potential toxicological risks of ENDS [22]. Their conclusion:

> “The use of E(N)NDS products, produced according to appropriate manufacturing standards and used as recommended, as a replacement for CC [cigarette] smoking, is likely to be associated with a reduction in overall risk of adverse health effects, although the magnitude of the decrease will depend on the effect in question.” (p. 28)

#### Biomarker Evidence

Except for one study on nicotine exposure, the *Opinion* does not present biomarker studies. Emissions testing is product testing, while biomarker studies provide evidence on actual human exposure to toxins. This is a serious shortcoming as research finds that exposure levels for a number of toxins are similar between ENDS users and non-smokers. Biomarker data is relevant evidence for demonstrating reductions in toxicant exposures from ENDS compared to cigarettes. The following studies represent some of the many biomarker studies that observe a substantial reduction in levels of toxins in the bodies of the participants who substituted ENDS for cigarette smoking.

The *Opinion* considered metal exposures, but did not cite a study [23] that reviewed blood lead (N=1899) and urinary cadmium, barium, and antimony (N=1302) test data in the 2015-2016 US National Health and Nutrition Examination Survey (NHANES). There were no significant differences in the levels of exposure to metals between participants who had never used ENDS and participants who were current or former ENDS users. The researchers conclude that ENDS are not a source of exposure to these heavy metals.

In a biomarker study funded by the US National Cancer Institute [24] 28 ENDS users who had quit smoking for a minimum of 2 months were given urine tests. The participants had significantly lower biomarkers of exposure compared to cigarette smokers. Their 1-HOP levels (exposure to polycyclic aromatic hydrocarbons) were similar to non-smokers. The study found significantly lower levels of metabolites in ENDS users compared to cigarette users. These were

* 1-hydroxypyrene/1-HOP (a marker for polycyclic aromatic hydrocarbons [PAH])

* 4-(methylnitrosamino)-1-(3-pyridyl)-1-butanol and its glucuronides/total NNAL (a marker for nicotine toxin)

* 3-hydroxypropylmercapturic acid/3-HPMA (a marker for acrolein)

* 2-hydroxypropylmercapturic acid/2-HPMA (a marker for propylene oxide)

* 3-hydroxy-1-methylpropylmercapturic acid/HMPMA (a marker for crotonaldehyde)

* *S*-phenylmercapturic acid/ SPMA (a marker for benzene).

The researchers conclude that

> “levels of a suite of urinary toxicant and carcinogen metabolites were significantly lower in e-cigarette users than in cigarette smokers. These results suggest that e-cigarette use may be safer than cigarette smoking, at least with respect to the compounds studied here, which represent typical carcinogens and toxicants believed to be involved in causing cancer in cigarette smokers.” (p. 708).

A clinical study [25] examined the effects of four weeks of ENDS substitution with 33 participants who smoked. The mean 3-HPMA levels (acrolein) decreased 79% for exclusive ENDS users and 60% for dual ENDS and cigarette users. Carbon monoxide levels decreased by 80% in exclusive ENDS users and 52% in dual users. Reduction of carbon monoxide levels to within normal limits has been reported soon after switching from conventional cigarette smoking to exclusive ENDS use in other studies [26-28].

A before and after study [29] tested 20 Polish adult cigarette users for biomarkers of exposure after 2 weeks, with half of the participants substituting ENDS and half continuing to smoke. Significant reductions in exposure levels were detected for many toxicants in the ENDS users, listed in Table 2 below.

**Table 2.** Significant Reductions in Toxin Exposures with Short-Term ENDS Substitution Source: [29, Supplemental Table 3]

| Toxicant | Significant Reduction<br>p<0.05 |
| --- | --- |
| NNK | 64% |
| Ethylene oxide | 61% |
| 1,3-Butadiene | 84% |
| Crotonaldehyde | 67% |
| Acrolein | 56% |
| Benzene | 76% |
| Acrylamide | 57% |
| Acrylonitrile | 79% |
| Propylene Oxide | 53% |
| PAH 1-Hydroxyfluorene | 58% |
| PAH 3-Hydroxyfluorene | 34% |

Source: [29, Supplemental Table 3]

The substantial and significant reduction in 1,3-butadiene is particularly noteworthy as it is the greatest source of cancer risk in tobacco smoke [30]. Goniewicz et al. conclude that “e-cigarettes may effectively reduce exposure to toxic and carcinogenic substances among smokers who switched to these products” (p. 165).

A 4-week observational study was conducted with 40 adult cigarette users who added or substituted ENDS use [31]. Biomarker levels for exposure to NNAL, benzene, and acrylonitrile were significantly reduced in all participants. Participants reporting exclusive ENDS use for at least two weeks exhibited additional significant reductions in metabolite levels of ethylene oxide and acrylamide. Exclusive ENDS users had reductions in acrolein levels bringing them into the range of non-smokers.

While significant reductions in biomarkers of exposure are not evidence of an absence of risk, these studies (and industry studies not cited) demonstrate that exposures to toxicants are substantially and significantly lower for ENDS users compared to those who smoke cigarettes. People who smoke can substantially reduce their exposure to known toxicants by substituting ENDS for cigarettes, even when it is not complete substitution.

#### Individual and Population Health Benefits

We are not aware of any studies showing a negative impact on the health of individuals who substitute ENDS for cigarette smoking. Although concern has been raised about the long-term health impacts of ENDS, a handful of short-term and long-term clinical trials have demonstrated health benefits from ENDS substitution. It is regrettable that there are so few clinical trials on ENDS health effects outside of cessation, although smoking cessation has known health benefits. Complete ENDS substitution for smoking may well prove over time to substantially lessen the risks of continued smoking.

In a 12-month assessment of the impact of ENDS use on blood pressure (BP) in 89 patients with hypertension, consistent and clinically significant improvement in systolic and diastolic BP as well as in BP control was observed in those who switched to regular ENDS use [32]. These findings are in agreement with the 8.8 mmHg reduction in systolic BP at 12-month in a prospective randomized control trial looking at the effect of smoking cessation by using ENDS in subjects with high BP at baseline [33].

A randomized controlled trial (N=114) [34] demonstrated that four weeks of ENDS substitution for smoking resulted in significant improvements in flow-mediated dilation and decreases in vascular stiffness compared to the cigarette user arm, indicating a reduced risk for cardiovascular disease.

The respiratory health effects of ENDS have been addressed in two recent review articles and their conclusions are conflicting [35, 36]. Both these reviews cite in vitro and in vivo studies, but findings from human studies support the view that ENDS use shows no evidence of health harms and even health improvements in patients with chronic obstructive pulmonary disease (COPD) or asthma.

In a small study of daily ENDS users who had never smoked, no noticeable changes in health outcomes were detected over the 3.5 year observation period [37]. Daily usage of ENDS caused no significant changes in measures of lung function, respiratory symptoms and lung inflammation and no significant structural abnormalities could be identified on high-resolution computerised tomography (HRCT) of the lungs.

A five year follow-up clinical study (assessments at 12, 24, 48, and 60 months) of patients with COPD compared 19 patients who completely or partially substituted ENDS for smoking to 20 controls who smoked [38]. Switching to ENDS contributed to a rapid improvement in cardiorespiratory health, with better quality of life, improved exercise tolerance, and an approximately 50% reduction in COPD exacerbations. Of note, these health gains were consistent throughout the 5-year follow-up.

A study [39] evaluated exclusive ENDS users with asthma who had stopped smoking. In one section of the study, a web survey (N=382), 91.6% self-reported no worsening of symptoms from ENDS use. The second part of the research conducted clinical testing of 10 ENDS users with asthma at baseline, 3 months, and 6 months. The participants experienced a significant increase in asthma symptom control and improvements in the Asthma Quality of Life Questionnaire (AQLQ) scores over the course of the study.

In a 2-year prospective clinical study, ENDS use consistently improved objective and subjective asthma outcomes [40]. ENDS use was well tolerated, and exposure to e-liquid aerosol in this vulnerable population did not trigger any asthma attacks.

A positive impact of ENDS substitution on population health has been also predicted by population-based models. Prochaska and Benowitz, leading expert researchers on tobacco, state

> “While e-cigarettes may have adverse effects on respiratory health and possibly other diseases, the harm is generally accepted to be much less than that of cigarette smoking. Thus, if smokers were to switch completely to e-cigarettes, then smoking-related disease is predicted to decrease substantially. Population-based models of the impact of e-cigarette use predict an overall health benefit” [41, pp. 17-18].

The WHO Study Group on Tobacco Product Regulation concurs, “The available evidence indicates a possible positive effect of ENDS on population health, particularly if appropriate ENDS regulation is enacted to maximize their benefits and minimize their risks” [10, p. 60]. The potential benefits of ENDS substitution for cigarette smoking on individual and population health certainly merit its inclusion in the final report.

### The Gateway Effect

#### Common Liabilities, an Alternative Hypothesis

The Terms of Reference specified an examination of a gateway effect. The gateway hypothesis is not the only explanation for the observed correlation between youth ENDS and cigarette use [9, 10]. The common liabilities theory posits that a “common latent propensity to risky behaviour” leads to concurrent cigarette and ENDS use [10, p. 57]. A review by the European Respiratory Society states that shared risk factors are “likely alternative explanations supported by the literature” [18, p. 14]. Hammond et al. [42] in a one year longitudinal cohort study (Canada, N=19,130) conclude that “it is highly plausible that ‘common factors’ account for a substantial proportion of increased cigarette-smoking initiation among e-cigarette users” (p. E1135). The common liabilities hypothesis that psychosocial factors explain poly-substance use by youth should be evaluated as an alternative explanation to a gateway effect.

In a recent editorial in the *American Journal of Public Health* [43], the gateway theory has been labelled as a “common deception.” Review teams are more sanguine, but unconvinced of a gateway hypothesis. A narrative review by [44] on youth ENDS use states that there is no strong evidence supporting the gateway hypothesis. A systematic review and meta-analysis [45] on adolescent ENDS use and smoking initiation reached the conclusion that “it is not clear how much of the relationship is causal (gateway effect) or is due to common liability…there is much less conclusive evidence for a gateway effect” (p. 13).

While no definitive explanation has been reached on why youth ENDS ever-users have a higher prevalence rate of smoking than those who never used ENDS, almost all of the studies come from US data. Considering this, we think it is important to note the evidence from a major survey of French youth conducted with compulsory participation for all 17 year olds [46]. In the 2017 survey (n=17,862 ever smokers) an analysis showed that youth who ever-used ENDS were less likely to be daily cigarette users (relative risk (RR)=0.62, CI 0.60-0.64) than ever-smokers who had never tried ENDS. Youth who tried ENDS first before ever-smoking were less likely to be daily cigarette users (RR=0.76, CI 0.66 – 0.89). Chyderiotis et al. observe that these findings are “in contradiction with the gateway hypothesis” (p. 5). Of course, no one study can prove or disprove a gateway effect, but this cross-sectional study is one of the very few studies available investigating a gateway effect among youth in an EU member state.

### Misleading Statements on EU Prevalence Trends

#### Renormalization

The *Opinion* alludes to a “resurgence of cigarette smoking” suggesting a renormalization of smoking. This is not happening, as clearly shown by the accelerated decline in smoking prevalence in EU member states, some of which have the highest prevalence of ENDS users. Changes in the trends of smoking prevalence are the best indicator of the absence or presence of renormalization [47]. Based on WHO data, between 2016 and 2018, 24 of 27 EU member states experienced declines in the prevalence of cigarette use for the population **15 years old and older**. Seven member states had cigarette prevalence declines of 6% or better and three member states had declines over 10% during the 2-year period. See Table 3 below.

**Table 3.**
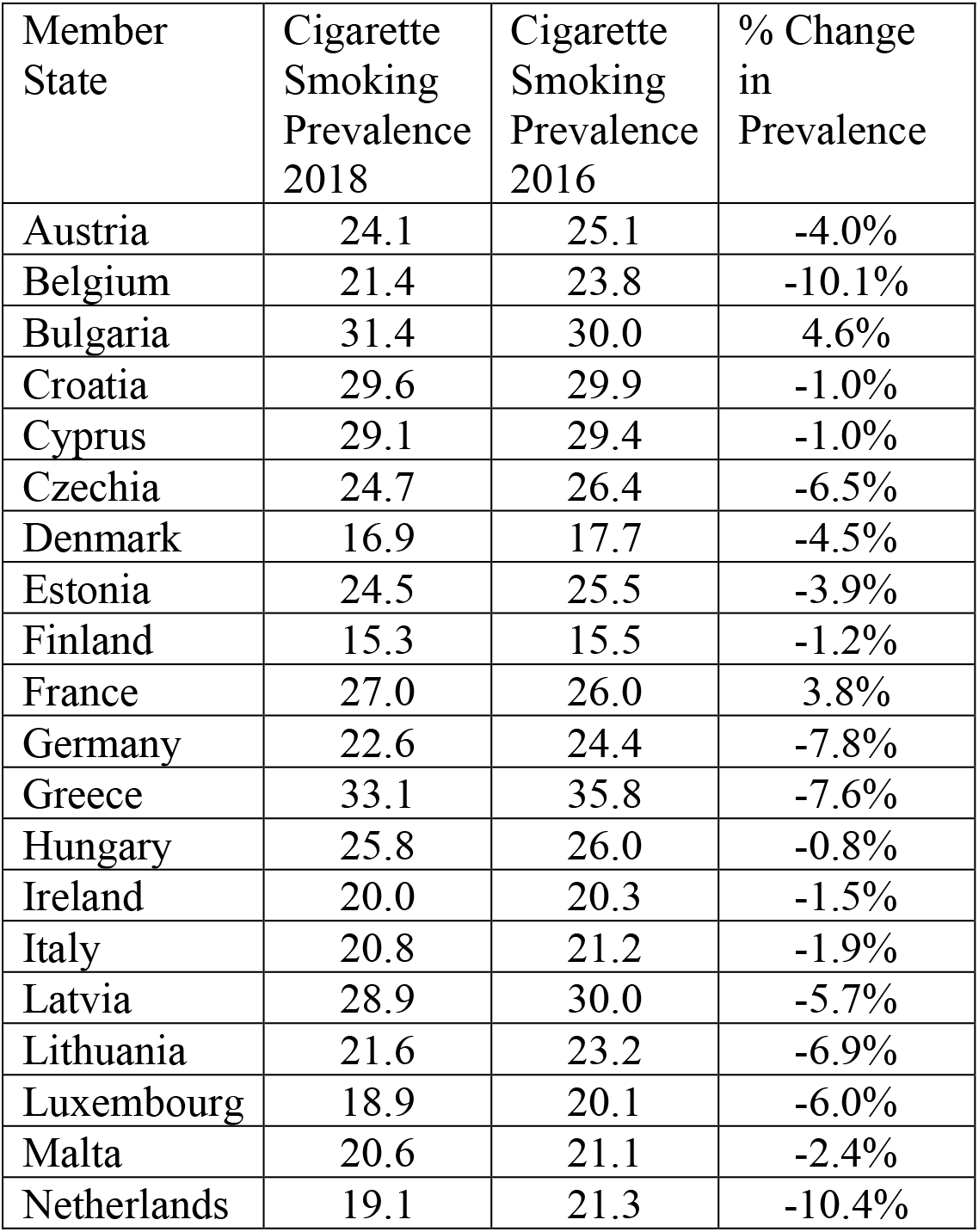
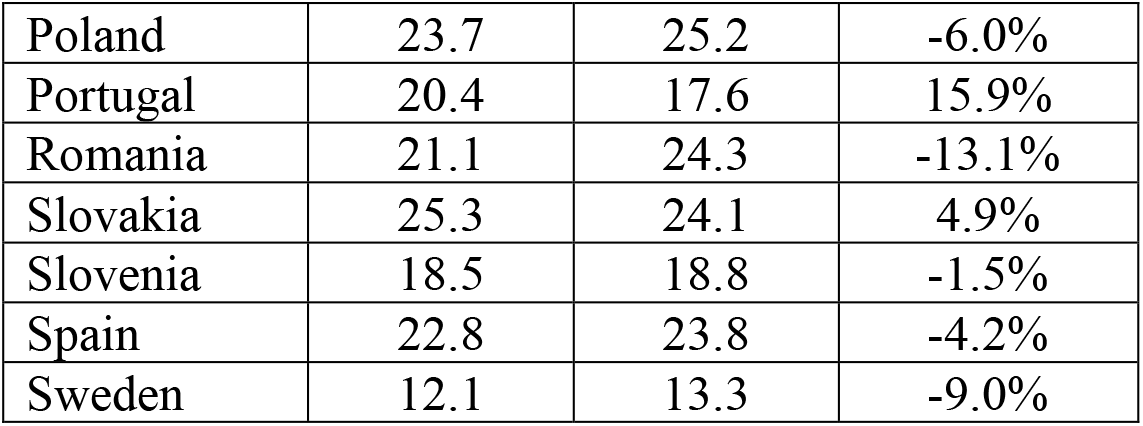
Declines/Increases in Cigarette Smoking Prevalence in EU Member States, 2016-2018 Source: World Health Organization [6, Table A1.4], World Health Organization [7, Table A.1.3]

| Member State | Cigarette Smoking Prevalence 2018 | Cigarette Smoking Prevalence 2016 | % Change in Prevalence |
| --- | --- | --- | --- |
| Austria | 24.1 | 25.1 | -4.0% |
| Belgium | 21.4 | 23.8 | -10.1% |
| Bulgaria | 31.4 | 30.0 | 4.6% |
| Croatia | 29.6 | 29.9 | -1.0% |
| Cyprus | 29.1 | 29.4 | -1.0% |
| Czechia | 24.7 | 26.4 | -6.5% |
| Denmark | 16.9 | 17.7 | -4.5% |
| Estonia | 24.5 | 25.5 | -3.9% |
| Finland | 15.3 | 15.5 | -1.2% |
| France | 27.0 | 26.0 | 3.8% |
| Germany | 22.6 | 24.4 | -7.8% |
| Greece | 33.1 | 35.8 | -7.6% |
| Hungary | 25.8 | 26.0 | -0.8% |
| Ireland | 20.0 | 20.3 | -1.5% |
| Italy | 20.8 | 21.2 | -1.9% |
| Latvia | 28.9 | 30.0 | -5.7% |
| Lithuania | 21.6 | 23.2 | -6.9% |
| Luxembourg | 18.9 | 20.1 | -6.0% |
| Malta | 20.6 | 21.1 | -2.4% |
| Netherlands | 19.1 | 21.3 | -10.4% |
| Poland | 23.7 | 25.2 | -6.0% |
| Portugal | 20.4 | 17.6 | 15.9% |
| Romania | 21.1 | 24.3 | -13.1% |
| Slovakia | 25.3 | 24.1 | 4.9% |
| Slovenia | 18.5 | 18.8 | -1.5% |
| Spain | 22.8 | 23.8 | -4.2% |
| Sweden | 12.1 | 13.3 | -9.0% |

#### ENDS Prevalence Trends

The highest ENDS prevalences are between 4.1% and 5.7% in eight member states while the prevalence is under 2% in 13 member states. Nor are EU prevalences “increasingly rising” as stated in the *Opinion*, and in fact they have been relatively stable from 2017 to 2019. During this period three member states had no increase in the prevalence and seven member states had an increase of 0.2 percentage points or less. Only two member states had an increase in the prevalence of 1.0% or higher, the Netherlands with 1.0% and Portugal with 1.3%. See Table 4 below.

**Table 4.** Prevalence of Adult ENDS use, EU Member States, 2017 and 2019 Source: [8 “Adult Smokers – Historical Data, Vapour Products”]

| Member State | ENDS prevalence 2019 | ENDS prevalence 2017 |
| --- | --- | --- |
| <u>Austria</u> | 1.6 | 0.9 |
| <u>Belgium</u> | 4.8 | 4.1 |
| <u>Bulgaria</u> | 1.1 | 1.1 |
| <u>Croatia</u> | 1.7 | 0.8 |
| <u>Cyprus</u> | NR | NR |
| <u>Czechia</u> | 5.7 | 5.6 |
| <u>Denmark</u> | 5.1 | 4.6 |
| <u>Estonia</u> | 1.6 | 1.9 |
| <u>Finland</u> | 2.0 | 1.9 |
| <u>France</u> | 4.4 | 4.2 |
| <u>Germany</u> | 5.4 | 5.2 |
| <u>Greece</u> | 2.6 | 2.4 |
| <u>Hungary</u> | 1.9 | 2.4 |
| <u>Ireland</u> | 5.5 | 5.3 |
| <u>Italy</u> | 1.7 | 1.4 |
| <u>Latvia</u> | 1.2 | 0.9 |
| <u>Lithuania</u> | 1.9 | 1.1 |
| <u>Luxembourg</u> | NR | NR |
| <u>Malta</u> | NR | NR |
| <u>Netherlands</u> | 4.1 | 3.1 |
| <u>Poland</u> | 5.4 | 5.4 |
| <u>Portugal</u> | 2.8 | 1.5 |
| <u>Romania</u> | 3.5 | 3.5 |
| <u>Slovakia</u> | 2.1 | 1.8 |
| <u>Slovenia</u> | 0.9 | 0.8 |
| <u>Spain</u> | 1.7 | 1.2 |
| <u>Sweden</u> | 1.0 | 1.0 |

NR = not reported
Source: [8 “Adult Smokers – Historical Data, Vapour Products”]

The Working Group cannot legitimately imply that ENDS may cause a resurgence of cigarette use because cigarette use had declined, often substantially, in almost every EU member state. Consumer data contradicts the statement that the prevalence of ENDS use among EU adults is rising quickly. In many EU member states ENDS usage prevalence has been relatively stable with only a very small rise, and in some member states no rise, between 2017 and 2019.

For data on the prevalence of youth use, the ESPAD report 2020, the *European School Survey Project on Alcohol and Other Drugs*, is due out November 12, 2020. Its findings should be included in the final report. Some prevalence data in the *Opinion* are from 2015 and earlier and so have a limited value for reporting current prevalences. Much of the prevalence data in the *Opinion* are for ever-use, a measurement that grossly over-represents the actual number of ENDS users, as discussed below.

### Mismeasurements in Exposure Assessments

#### Frequency of Use

The *Opinion* frequently cites ever-use data as if it were evidence of usage prevalence. Ever-use is a problematic measurement that captures a substantial number of one-time triers and substantially overestimates actual usage. Indeed, it does not even track it, given that within a cohort ever-use will always increase over time even if usage prevalence drops.

Ever-use data on youth users is unreliable for exposure assessment “as ever use can include using an e-cigarette once across the lifetime, the extent of increased nicotine exposure as a result of ever e-cigarette use is unclear” [48, p. 616]. Data from the Global Youth Tobacco Survey (GYTS) shows that 27% to 55% (varying by country) of EU youth who ever tried ENDS did so on only one occasion.

For Italian youth (15-19 years old), the ESPAD Italy 2017 survey found that over 70% of youth ever-users had used ENDS only one to nine times [50, supplementary material].

Ever-use measurement counts a substantial number of youth ENDS experimenters who do not go on to become regular users [51]. Similarly, among youth aged 13 to 17 in Norway in a four year longitudinal (2015-2019) qualitative study (50 semi-structured group and 175 individual interviews), it was often the case that as youth became older, ENDS use became viewed as a childish practice that they discarded [52].

Current youth use, defined as any use in the past 30 days, includes a substantial number of youths who use ENDS on only one or two days, including those who only tried ENDS once in their lives but it happened to be during that month. See Table 6 below.

**Table 5.**
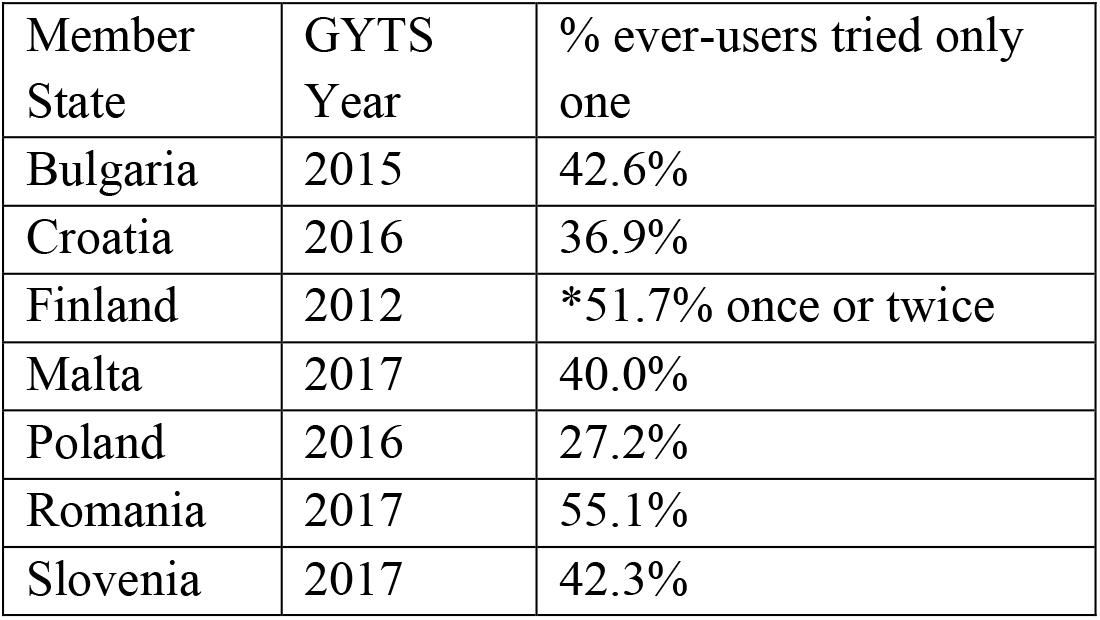
Prevalence of One Occasion ENDS Use in Youth Ever-Users Source: World Health Organization [49] “Electronic cigarette smoked all life”

| Member State | GYTS Year | % ever-users tried only one |
| --- | --- | --- |
| Bulgaria | 2015 | 42.6% |
| Croatia | 2016 | 36.9% |
| Finland | 2012 | *51.7% once or twice |
| Malta | 2017 | 40.0% |
| Poland | 2016 | 27.2% |
| Romania | 2017 | 55.1% |
| Slovenia | 2017 | 42.3% |

**Table 6.** Prevalence of Infrequent ENDS Use Among Past-30-Day Youth Users Source: World Health Organization [49] “Electronic cigarettes past month”

| Member State | GYTS Survey Year | % past month use of 1-2 days |
| --- | --- | --- |
| Bulgaria | 2015 | 51.6% |
| Croatia | 2016 | 61.6% |
| Czech | 2016 | 46.2% |
| Finland | 2012 | *61.8% |
| Italy | 2018 | 59.2% |
| Latvia | 2014 | 58.3% |
| Malta | 2017 | 47.0% |
| Poland | 2016 | 42.2% |
| Romania | 2017 | 55.9% |
| Slovakia | 2016 | 56.1% |

\* less than once a week

Ever-use data for EU adults is also problematic for exposure assessment and estimating prevalence. The 2016 European Regulatory Science on Tobacco (EUREST-PLUS ITC, N=1178) found that among adult ever-users in Germany, Greece, Hungary, Poland, Romania, Spain, 38% had used ENDS 1-2 times and 21% had used ENDS 3-10 times. Furthermore 85% of ever-users were no longer using ENDS [53]. Current use of ENDS defined as any-past-30-day use is not a good indicator of sustained use as infrequent users of five days or less a month “includes many individuals who can be expected to discontinue use within 1 year” [54, p. e92].

The measurement of ever-use for youth and adults results in a misrepresentation of prevalence because many are simply experimenting on a very limited basis and do not continue use. The measurement of any-past-30-day use, the classification for a current user, implies a regular pattern of use. This representation is belied by the finding that 42% - 61% of EU youth defined as current users are infrequent users, only once or twice a month. This measurement may also be capturing recent triers.

#### Non-Nicotine Use

The *Opinion* makes no reference to use of non-nicotine ENDS, an omission of important data on nicotine exposure. In the EU, evidence shows that a substantial number of youth and adults do not use nicotine liquids.

Many EU youth ENDS ever-users report that they used non-nicotine products. In Finland: 52% of boys and 48% of girls used only non-nicotine ENDS, and nicotine use declined from 2013 to 2019 [55]. In France: 42.2% of ever-smokers and 92.9% of non-smokers used only non-nicotine ENDS [56]. In Italy: 72.0% overall, 40.3% used only non-nicotine and 31.7% used both nicotine and non-nicotine [49]. In Sweden: 38% used only non-nicotine ENDS [57]. A two year longitudinal cohort study in Finland (N=3474) found that exclusive use of non-nicotine ENDS by adolescents was not associated with a higher probability of becoming a daily smoker compared to never ENDS users [58].

Non-nicotine ENDS use also appears to be common among youth classified as “current” (recent) ENDS users. A US survey (N=1589, 15-17 years old) reporting past-30-day use found 29% used non-nicotine liquids and 39% reported using both nicotine and non-nicotine liquids [59].

Three studies indicate that many EU adults ENDS users are using non-nicotine liquids. A 2016 survey of French young adults (19-22 years old) recent ENDS users found 61 of 98 used only non-nicotine ENDS and an additional 19 reported using both [60]. A 2016 face-to-face interview project with 600 adult daily ENDS users in Barcelona, Spain reported that 33.7% of users quitting smoking and 43.6% of users reducing cigarette use did not use nicotine liquids [61]. In a 2016 online survey of current ENDS users in Poland (N=1142), 9.8% started ENDS use because they could use non-nicotine liquids [62].

Nicotine use during pregnancy is a known risk factor for adverse neonatal outcomes. A 2015 survey [63] of women in two US states having given birth that year found that 35.2% of women who used ENDS in the last three months of pregnancy (even once) used non-nicotine ENDS. During the period of three months before pregnancy to 2–6 months after delivery, 41.4% of ever-users used only non-nicotine ENDS.

Exposure assessments for ENDS use are complex. Approximately 75% of the *Opinion’s* statements on exposures are based on ever-use data. This data should either be excluded in the final report, or flagged to be as viewed with caution, because most of those included in these counts either used ENDS only a very small number of times or exclusively used non-nicotine ENDS. Infrequent use should, at a minimum, be noted in the exposure assessments. Reporting the prevalence of non-nicotine use is critical to exposure assessments because of its implications for the risks of nicotine addiction, pregnancy outcomes, and for the cardiovascular effects of ENDS use (see section following).

### Incorrect Assessment of Cardiovascular Risk

The *Opinion* states that there is strong evidence for long-term systematic effects of ENDS on the cardiovascular system. This is an egregious error. The statement on cardiovascular risk is contradicted by numerous researchers. The National Academies of Sciences, Engineering, and Medicine systematic review [9] concludes that “There is **no available evidence** whether or not e-cigarette use is associated with clinical cardiovascular outcomes (coronary heart disease, stroke, and peripheral artery disease) and subclinical atherosclerosis (carotid intima-media thickness and coronary artery calcification)” (p.7, emphasis in original). Benowitz and Fraiman [64] and D’Amario et al. [65] state that there is no available evidence on cardiovascular risk. Münzel et al. [66] in their review conclude that strong evidence on long term effects is missing.

Two review teams observe that the assessment of cardiovascular risk is controversial, and that any risk may be attributed solely to nicotine [10, 67]. This controversy is evidenced in the conflicting results from two clinical studies. A randomized cross-over trial (N=25) of nicotine and non-nicotine ENDS use did not find negative changes in micro and macrovascular endothelial function or oxidative stress with non-nicotine ENDS [68]. This group of researchers conclude that negative the cardiovascular effects of ENDS use are attributable to the nicotine present in the liquid. Yet the clinical trial by George et al. [34] discussed earlier found no difference in cardiovascular effects between nicotine and non-nicotine ENDS.

Benowitz and Burbank [69] characterized the cardiovascular risks of ENDS as “very low” for those with no known cardiovascular disease, while those with cardiovascular disease “might incur some increased risk” albeit far lower than smoking (p. 521). They conclude that “If e-cigarettes can be substituted completely for conventional cigarettes, the harms from smoking would be substantially reduced and there would likely be a substantial net benefit for cardiovascular health” (p. 521).

A four month randomized observational study divided 40 participants who smoked into an ENDS arm and a continued smoking arm [70]. The ENDS participants demonstrated reductions in arterial stiffness and oxidative stress that was not experienced by the participants who continued to smoke.

A large US dataset, the National Health Interview Survey, was examined for evidence of ENDS use and cardiovascular risks by [71]. A pooled analysis of the 2016 and 2017 surveys did not find an association between ENDS use and myocardial infarction or coronary heart disease.

The *Opinion’s* statement that the overall weight of the evidence of ENDS use for long-term systemic cardiovascular effects is strong is an overstatement. Consequently, it deserves to be re-evaluated and revised in the final report.

### Misrepresentation on the Role of Flavours

Contrary to the picture presented in the *Opinion*, the most common reason by far for youth and young adult ENDS experimentation is curiosity, not flavours. A 2018 survey of French youth (age 15-16, N=1435) found that that curiosity was the most common reason for trying ENDS, followed by flavours [72, data not reported]. A 2016 survey of ever-users in Germany (n=474) aged 14 and older reported that 59.5% endorsed curiosity as the reason for trying ENDS, increasing to 73.1% in the 14-19 year old age group [73]. For French young adults (19-22 years old) a 2016 survey (N=2720) found that 77.4% tried ENDS out of curiosity, 63.5% because someone offered it to them, and 24.6% because of flavours [60]. In the 2019 US National Youth Tobacco Survey (NYTS) [74] flavours ranked third in the reasons for use (22.3%), with curiosity the major reason for trying ENDS (56.1%), followed by use by family or peers (23.9%).

Interestingly, the ability to use ENDS for playing tricks was just as common a reason as flavours for trying ENDS at 21.2% in the 2019 NYTS. Tricks with ENDS are very popular among US youth. In a 2017 survey (N=2945 high school students) 54.4% of ever-users had done vape tricks [75]. Approximately three out of four US youth past-30-day ENDS users said they performed vape tricks; in a 2016 survey the number was 75% (N=1,729, aged 15-17) [76] and 73% of current users another survey [75]. It appears that vape tricks may attract adults too. In a small survey (n=183 ENDS users) conducted in the Netherlands 24.6% endorsed cloud chasing tricks as an attractive feature of ENDS [77]. Attractive features, such as the generation of very dense aerosols, appear to entice people into using these products and not just flavors.

The use of flavours does not appear to increase the risk of cigarette initiation for youth. A US longitudinal cohort study [78] analyzed data from waves 1 to 4 of the Population Assessment of Tobacco and Health (PATH) Study (2013 to 2018, n=7311) and the use of ENDS with nontobacco flavours was less strongly associated with youth smoking initiation than the use of tobacco flavours (AOR 0.66; CI 0.16-2.76).

The use of flavours appears to improve cessation success rates for adults. The PATH longitudinal cohort study noted above [78] also analyzed cessation by adults under 55 years old (n=5984), and the use of non-tobacco flavours was more strongly associated with smoking cessation compared to tobacco flavours (2.28; CI 1.04-5.01). This finding is corroborated by the longitudinal data analysis of PATH waves 1 through 3 by another research group [79]. Their analysis broke down the increase in quit rates by length of cessation, with non-tobacco flavoured ENDS substantially associated with higher quit rates among US adults using ENDS for smoking cessation, RRR 1.75 (CI 1.18-2.60) for past year quitters and RRR 2.83 (CI 1.69-4.73) for 1+ year cessation.

### Cessation

A large number of respondents in an EU survey report using ENDS in their smoking quit attempt, averaging about one in five.

The use of ENDS for quitting is widespread in France, with 76.3% of those who had quit smoking for at least one month stating they had used ENDS as a quit aid [2017 Eurobarometer Survey, 80].

Evidence presented in three reviews points to ENDS as an effective cessation aid for those who are already attempting to quit smoking. The most recent Cochrane review [81], published subsequent to the posting of the *Opinion*, concludes there is moderate-certainty evidence that ENDS use for cessation results in a higher quit rate than NRT, RR 1.69 (CI 1.25-2.27). ENDS produced a higher quit rate than behavioural support only or no support, RR 2.50 (CI 1.24-5.04), although the evidence is of very low certainty. A systematic review and meta-analysis [82] of 14 studies with 35,665 participants calculated quit rate efficiency from 13.2% to 22.9%. The reviewers characterize ENDS as a “promising” cessation aid. A review [83] in *Pharmacotherapy*, a journal of the American College of Clinical Pharmacy, states that ENDS “may have modest effects to help tobacco users achieve cessation” in a number of different patient populations (p. 565). In addition to these reviews, longitudinal data from the US PATH surveys (n=9724) showed that people making a quit attempt with ENDS were 1.32 (CI 1.03-1.71) times more likely to quit in the past year than those making a quit attempt without ENDS [79].

At the clinical level, a Belgium case report of ENDS use for cessation by patients in treatment with tobacco counselors, at 7 months (n=103, 70 ENDS users) almost 40% had biochemical verified abstinence, RR 1.71 (CI 1.04-2.81) compared to nicotine replacement therapy users [84]. ENDS are recommended as cessation help by the UK National Health Services on its website: *Using e-cigarettes to stop smoking*. It states that “Many thousands of people in the UK have already stopped smoking with the help of an e-cigarette. There’s growing evidence that they can be effective” [85].

### *Preliminary Opinion* References

Of note, we identified 21 articles in the References that are not cited in the text and provided this information to the Committee in our submission. Listing these articles as references without any discussion misrepresents which studies were evaluated or how they were interpreted by the Committee.

### Conclusions

Based on the evidence presented, the following changes are recommended to the Abstract for the final report:

1. There is little or no evidence on cardiovascular risks from ENDS use.
2. The evidence that supposedly supports claims of a gateway effect is consistent with the inevitable association between the propensity to use both ENDS and cigarettes.
3. A statement should be added that a substantial number of youth and adults use non-nicotine liquids.
4. Curiosity is the primary reason for youth experimentation.
5. The evidence for ENDS as an effective aid in quit attempts is <u>moderate</u>. Furthermore, the Abstract should include a statement on ENDS substitution for cigarette smoking. The SCHEER mandate for the scientific opinion included tobacco harm reduction, therefore it should be included in the Abstract of the final report.

The Summary section of the final report should include these areas: a detailed presentation on ENDS use for cigarette substitution, a discussion of alternative explanations and contradictory evidence for a gateway effect, an explanation of caveats for the measurement of frequency of use, an overview of all the factors for ENDS experimentation by youth, and a more detailed discussion of ENDS use for cessation.

The point of this critique is not to denigrate the *Preliminary Opinion*, but to rectify its errors and omissions in the final report. We hope that the literature cited here will prove useful for inclusion in the final report. Our goal is to support the revision of the *Preliminary Opinion* into a more robust and factually accurate final report. This is no minor matter – the lives and health of millions of European Union citizens are at stake.

## Data Availability

Data for Tables 1, 2, 3, and 7 were extracted from the published articles cited as the source.
Data for Table 4 is from the Passport database and is not publicly available. It is available only by subscription from EUROMONITOR International, London, a privately owned market research company. The company can be contacted at https://www.euromonitor.com/
Data for Tables 5 and 6 are from the Global Youth Tobacco Surveys with open access at the World Health Organization NCD Microdata Repository: https://extranet.who.int/ncdsmicrodata/index.php/home.

https://www.euromonitor.com/

https://extranet.who.int/ncdsmicrodata/index.php/home

## List of Abbreviations

BP: blood pressure
CI 95%: confidence interval
COPD: chronic obstructive pulmonary disease ENDS electronic nicotine delivery system(s)
ESPAD: *European School Survey Project on Alcohol and Other Drugs*
EU: European Union
GYTS: Global Youth Tobacco Survey
N: total number of participants
n: subset of participants NR not reported
NYTS US: National Youth Tobacco Survey
PATH US: Population Assessment of Tobacco and Health Survey RR relative risk
RRR: relative risk ratio
US: United States
WHO: World Health Organization

## Declarations

### Ethics approval and consent to participate

Not applicable.

### Availability of data and materials

Data for Tables 1, 2, 3, and 7 were extracted from the published articles cited as the source.

**Table 7.**
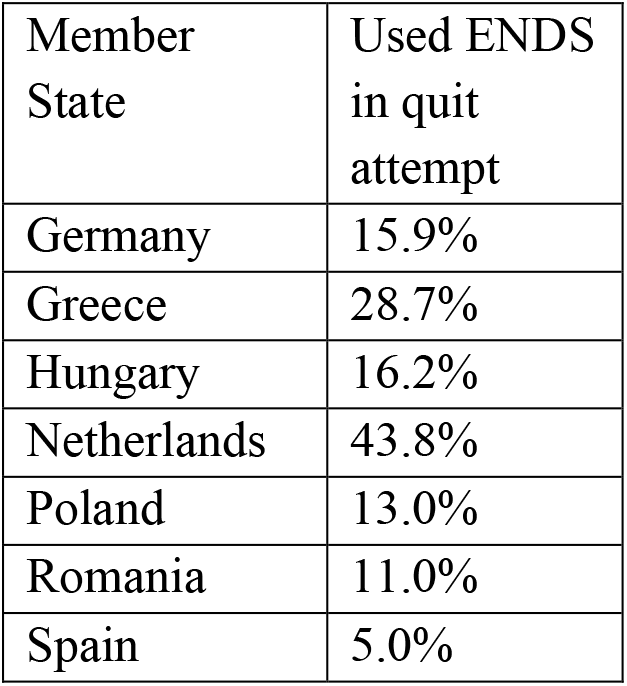
Use of ENDS for a Quit Attempt, EU Member States Source: [13]

Data for Table 4 is from the Passport database and is not publicly available. It is available only by subscription from EUROMONITOR International, London, a privately owned market research company. The company can be contacted at https://www.euromonitor.com/

Data for Tables 5 and 6 are from the Global Youth Tobacco Surveys with open access at the World Health Organization NCD Microdata Repository: https://extranet.who.int/ncdsmicrodata/index.php/home.

## Authors’ Contributions

All authors contributed to the conception of the work. RO conducted the literature search and RP submitted articles for inclusion. RO conducted the data extractions and analyses for the Tables. RO and RP performed the analysis and drafted the article. All authors submitted revisions to the draft for the final manuscript. All authors have approved the submitted version and are personally accountable for the accuracy and integrity of the entire work, as well as any modified version submitted after peer review.

## Acknowledgements

Members of the CoEHAR study group (listed alphabetically):

1. Salvatore **ALAIMO**, Department of Clinical and Experimental Medicine, University of Catania, Italy
2. Carmelina Daniela **ANFUSO**, Department of Biomedical and Biotechnological Sciences, University of Catania, Italy
3. Ignazio **BARBAGALLO**, Department of Drug Sciences, University of Catania, Italy
4. Francesco **BASILE**, Department of General Surgery and Medical-Surgical Specialties, University of Catania, Italy
5. Sebastiano **BATTIATO**, Department of Mathematics and Computer Sciences, University of Catania, Italy
6. Gaetano **BERTINO**, Department of Clinical and Experimental Medicine, University of Catania, Italy
7. Alberto **BIANCHI**, Department of General Surgery and Medical-Surgical Specialties, University of Catania, Italy
8. Antonio G. **BIONDI**, Department of General Surgery and Medical-Surgical Specialties, University of Catania, Italy
9. Maria Luisa **BRANDI**, National Observatory of Fragility Fractures, Italy
10. Emma **CACCIOLA**, Department of Medical, Surgical Sciences and Advanced Technologies, University of Catania, Italy
11. Rossella R. **CACCIOLA**, Department of Clinical and Experimental Medicine, University of Catania, Italy
12. Bruno Santi **CACOPARDO**, Department of Clinical and Experimental Medicine, University of Catania, Italy
13. Aldo E. **CALOGERO**, Department of Clinical and Experimental Medicine, University of Catania, Italy
14. Maria Teresa **CAMBRIA**, Department of Biological, Geological and Environmental Sciences, University of Catania, Italy
15. Davide **CAMPAGNA**, Department of Emergency Medicine, University of Catania Teaching Hospital Policlinico, Italy
16. Filippo **CARACI**, Department of Drug Sciences, University of Catania, Italy
17. Agatino **CARIOLA**, Department of Law Sciences, University of Catania, Italy
18. Massimo **CARUSO**, Department of Biomedical and Biotechnological Sciences, University of Catania, Italy
19. Pasquale **CAPONNETTO**, Department of Educational Sciences, University of Catania, Italy
20. Fabio **CIBELLA**, Institute of Biomedicine and Molecular Immunology, National Research Council, Italy
21. Maurizio **DI MAURO**, Department of Clinical and Experimental Medicine, University of Catania, Italy
22. Santo **DI NUOVO**, Department of Educational Sciences, University of Catania, Italy
23. Adriana **DI STEFANO**, Department of Law Sciences, University of Catania, Italy
24. Filippo **DRAGO**, Department of Biomedical and Biotechnological Sciences, University of Catania, Italy
25. Salvatore **FAILLA**, Department of Chemical Sciences, University of Catania, Italy
26. Rosario **FARACI**, Department of Economics and Business, University of Catania, Italy
27. Salvatore **FERLITO**, Department of Medical, Surgical Sciences and Advanced Technologies, University of Catania, Italy
28. Margherita **FERRANTE**, Department of Medical, Surgical Sciences and Advanced Technologies, University of Catania, Italy
29. Alfredo **FERRO**, Department of Clinical and Experimental Medicine, University of Catania, Italy
30. Giancarlo A. **FERRO**, Department of Law Sciences, University of Catania, Italy
31. Francesco **FRASCA**, Department of Clinical and Experimental Medicine, University of Catania, Italy
32. Lucia **FRITTITTA**, Department of Clinical and Experimental Medicine, University of Catania, Italy
33. Pio M. **FURNERI**, Department of Biomedical and Biotechnological Sciences, University of Catania, Italy
34. Antonio **GAGLIANO**, Department of Electrical, Electronics and Computer Engineering, University of Catania, Italy
35. Giovanni **GALLO**, Department of Mathematics and Computer Sciences, University of Catania, Italy
36. Fabio **GALVANO**, Department of Biomedical and Biotechnological Sciences, University of Catania, Italy
37. Giuseppe **GRASSO**, Department of Chemical Sciences, University of Catania, Italy
38. Francesca **GUARINO**, Department of Biomedical and Biotechnological Sciences, University of Catania, Italy
39. Antonino **GULINO**, Department of Chemical Sciences, University of Catania, Italy
40. Emmanuele A. **JANNINI**, Department of Systems Medicine, University of Rome Tor Vergata
41. Sandro **LA VIGNERA**, Department of Clinical and Experimental Medicine, University of Catania, Italy
42. Giuseppe **LAZZARINO**, Department of Biomedical and Biotechnological Sciences, University of Catania, Italy
43. Antonio **LONGO**, Department of General Surgery and Medical-Surgical Specialties, University of Catania, Italy
44. Gabriella **LUPO**, Department of Biomedical and Biotechnological Sciences, University of Catania, Italy
45. Mario **MALERBA**, Department of Translational Biomedicine, University of Eastern Piedmont, Italy
46. Luigi **MARLETTA**, Department of Electrical, Electronics and Computer Engineering, University of Catania, Italy
47. Guido **NICOLOSI**, Department of Political and Social Sciences, University of Catania, Italy
48. Francesco **NOCERA**, Department of Electrical, Electronics and Computer Engineering, University of Catania, Italy
49. Gea **OLIVERI CONTI**, Department of Medical, Surgical Sciences and Advanced Technologies, University of Catania, Italy
50. Rosalba **PARENTI**, Department of Biomedical and Biotechnological Sciences, University of Catania, Italy
51. Alfredo **PULVIRENTI**, Department of Clinical and Experimental Medicine, University of Catania, Italy
52. Francesco **PURRELLO**, Department of Clinical and Experimental Medicine, University of Catania, Italy
53. Francesco **RAPISARDA**, Department of Clinical and Experimental Medicine, University of Catania, Italy
54. Venerando **RAPISARDA**, Department of Clinical and Experimental Medicine, University of Catania, Italy
55. Michele **REIBALDI**, Department of General Surgery and Medical-Surgical Specialties, University of Catania, Italy
56. Renata **RIZZO**, Department of Clinical and Experimental Medicine, University of Catania, Italy
57. Simone **RONSISVALLE**, Department of Drug Sciences, University of Catania, Italy
58. Martino **RUGGIERI**, Department of Clinical and Experimental Medicine, University of Catania, Italy
59. Maria C. **SANTAGATI**, Department of Biomedical and Biotechnological Sciences, University of Catania, Italy
60. Cristina **SATRIANO**, Department of Chemical Sciences, University of Catania, Italy
61. Laura **SCIACCA**, Department of Clinical and Experimental Medicine, University of Catania, Italy
62. Maria Salvina **SIGNORELLI**, Department of Clinical and Experimental Medicine, University of Catania, Italy
63. Marco **TATULLO**, Technologica Research Institute, Marrelli Hospital, Italy
64. Daniele **TIBULLO**, Department of Biomedical and Biotechnological Sciences, University of Catania, Italy
65. Venera **TOMASELLI**, Department of Political and Social Sciences, University of Catania, Italy
66. Luca **ZANOLI**, Department of Clinical and Experimental Medicine, University of Catania, Italy
67. Agata **ZAPPALÀ**, Department of Biomedical and Biotechnological Sciences, University of Catania, Italy

